# Food supplementation and behavioral counseling to reduce the double burden of malnutrition among pregnant women and their infants: protocol for the K’ASLEM hybrid type 1 effectiveness-implementation randomized controlled trial

**DOI:** 10.64898/2026.09.08.26362539

**Authors:** David Flood, Mónica Mazariegos, Sara Hernández Hidalgo, Yolanda Juarez Martin, Wendy Azucena Rodríguez Gómez, Michelle H. Moniz, Chunling Lu, Ann C. Miller, Manuel Ramírez-Zea, Peter Rohloff

## Abstract

**Introduction:** Adults globally are experiencing increases in diseases attributable to overnutrition, yet child undernutrition persists at high levels. This “double burden of malnutrition” increases the risk of nutrition-related non-communicable diseases in mother and child. Evidence-based interventions promote optimal maternal weight or child growth, but gaps remain in implementing them as integrated, sustainable, and equity-focused solutions.

**Methods and analysis:** K’ASLEM is an individually randomized, parallel-group Hybrid Type 1 effectiveness-implementation trial in Indigenous Maya communities in Guatemala with high levels of the double burden of malnutrition. We will enroll 766 pregnant women aged ≥16 years at less than 28 weeks’ gestation, allocated 1:1 to intervention or comparator. The intervention integrates (a) food supplementation for pregnant and postpartum mothers and their infants and (b) behavioral counseling to optimize maternal weight and promote healthy nutrition, physical activity, and infant feeding practices. The comparator is enhanced usual care. Mother-child dyads will be followed until 12 months after birth. Co-primary outcomes are maternal weight and child length-for-age z-score at 12 months. Secondary outcomes are child stunting, child development, maternal overweight or obesity, and maternal hemoglobin. We will collect quantitative and qualitative implementation data, use Implementation Mapping to identify factors shaping implementation, and conduct an economic evaluation of costs and cost-effectiveness.

**Ethics and dissemination:** Ethics approval was obtained from the Institutional Review Boards of Mass General Brigham and Maya Health Alliance and the Institute of Nutrition of Central America and Panama Research Ethics Committee. Results will be published in peer-reviewed, open-access journals.

**Trial registration:** This trial was prospectively registered on ClinicalTrials.gov (NCT06750120).

**Protocol version:** Version 3.0 (March 17, 2026).

**KEY MESSAGES:** *What is already known on this topic:* The double burden of malnutrition, commonly defined at the household level as maternal overweight or obesity and child stunting in a mother-child dyad, occurs in more than 20% of households in some low- and middle-income countries. Inadequate nutrition during pregnancy, the postpartum period, and early childhood increases the lifelong risk of nutrition-related non-communicable diseases in both mother and child. Evidence-based interventions promote optimal maternal weight and child growth, but critical knowledge gaps remain in implementing them as integrated, sustainable, and accessible solutions among the world’s poorest.

*What this study hopes to add:* The K’ASLEM randomized controlled trial addresses these gaps by assessing an integrated intervention to reduce the double burden of malnutrition among pregnant and postpartum women and their children in Guatemala. The intervention consists of (1) food supplementation and (2) behavioral counseling to optimize maternal weight during pregnancy and the postpartum period and to promote healthy nutrition, physical activity, and infant feeding practices.

*How this study might affect research, practice or policy:* This study will generate evidence on the effectiveness, implementation, and cost-effectiveness of an integrated intervention to address the double burden of malnutrition, with implications for nutrition and non-communicable disease policy in Guatemala and other countries.

## INTRODUCTION

Adult populations globally are experiencing rapid increases in non-communicable diseases (NCDs) attributable to overnutrition, yet child undernutrition persists at extraordinary levels.^1^ This “double burden of malnutrition” (DBM) occurs at the population, household, and individual levels.^2^ At the household level, the focus of this trial, the DBM is commonly conceptualized as the coexistence within a mother-child dyad of excess maternal weight and impaired child linear growth.^1^ Classified by the dichotomous thresholds used in epidemiological monitoring (maternal body mass index ≥25 kg/m2 and child length-for-age z-score <-2), the household-level DBM affects more than 20% of households in some low- and middle-income countries and is disproportionately concentrated among socioeconomically disadvantaged households.^3^ Pregnancy, postpartum, and early childhood are critical life stages in which to address the DBM. Among children, linear growth failure during these stages is associated with profound biological, developmental, and economic consequences throughout the lifespan, including an increased risk of NCDs.^4^ Among mothers, excess gestational weight gain and postpartum weight retention increase risk of pregnancy complications, NCDs, and mortality.^5^

Addressing the DBM requires “double-duty actions,” or interventions that target undernutrition and overnutrition together rather than as separate problems.^6^ There are evidence-based interventions that prevent child growth failure or promote optimal maternal weight, but critical knowledge gaps remain in implementing them as integrated, sustainable, and accessible solutions among the world’s poorest.^6^ Two interventions with strong evidence are (1) food supplementation during pregnancy and the postpartum period for mothers and upon the transition to complementary foods for infants;^7,8^ and (2) behavioral counseling to optimize maternal gestational weight gain and limit postpartum weight retention.^9-11^

Despite the proven efficacy of these interventions in isolation, their population benefits have been limited for a few key reasons. Food supplementation improves child growth but may cause excessive maternal weight gain during and after pregnancy.^6,12^ Implementation knowledge gaps also have limited the scale-up of effective nutrition interventions. Efficacy trials of nutrition supplementation have generally paid limited attention to implementation or sustainability, and behavioral counseling trials among pregnant or postpartum women have been conducted almost exclusively in high-income countries. Prior trials have often excluded the populations most affected by the DBM, which limits generalizability among the most at-risk households.

The K’ASLEM trial attempts to address these research gaps by assessing the effectiveness, implementation, and cost-effectiveness of an integrated intervention to reduce the double burden of malnutrition among pregnant and postpartum women and their children in Guatemala.

## METHODS AND ANALYSIS

This protocol follows the SPIRIT guidelines for clinical trial protocols.^13^ We draw on applicable items from the Standards for Reporting Implementation Studies (StaRI) checklist to guide reporting of the implementation research component of this protocol.^14^

### Study design

The K’ASLEM trial is an individually randomized, parallel-group Hybrid Type 1 effectiveness-implementation clinical trial conducted in rural Guatemala. The word “k’aslem,” which means “life” in the Kaqchikel Mayan language, was chosen to reflect the holistic conceptualization of wellbeing in the Maya cosmovision in which physical health, nourishment, and new life are interconnected. A Hybrid Type 1 design is appropriate because the intervention has strong face validity and low risk, but limited evidence for effectiveness in this context.^15^ Recruitment began in May 2026, and final data collection is anticipated in June 2029.

### Study setting

The K’ASLEM trial will be carried out in majority Indigenous Maya municipalities in the department of Chimaltenango, Guatemala. The government’s national nutrition plan prioritizes the double burden of malnutrition and designates trial municipalities as among the priority geographies for both child undernutrition and maternal overweight and obesity.^16^ The national prevalence of household-level DBM in Guatemala is approximately 20-25%,^17,18^ among the highest in the world,^3^ and it is even higher in these municipalities. Approximately 70% of Indigenous Guatemalan children under five years of age are stunted,^19^ and about half of non-pregnant reproductive-aged women in the study setting are overweight or obese.^19^ Slightly more than half of Maya households in this area live below the poverty line, defined as the per-capita cost of minimum caloric and non-food needs.^20^

Maya Health Alliance and the Institute of Nutrition of Central America and Panama (INCAP) will co-lead the trial. Maya Health Alliance is a non-governmental organization specializing in health care and research in Indigenous Maya communities in Guatemala. INCAP is a publicly governed research institution whose mission is to provide technical support to improve nutrition policy in Guatemala and other countries in Central America.

### Eligibility criteria

#### Inclusion criteria

1. Women aged 16 years or older.
2. Gestational age less than 28 weeks.

#### Exclusion criteria

1. History of pregestational diabetes (type 1 diabetes or type 2 diabetes), history of gestational diabetes in a previous pregnancy, or diagnosis of gestational diabetes in the current pregnancy.
2. Multifetal gestation (twins or higher-order pregnancies).
3. Currently participating in another research study involving an intervention.
4. Has a family member who has already been invited to participate in this study and/or shares a kitchen with such a person.
5. Has a serious underlying medical or psychiatric condition requiring specialized clinical care, including active cancer, severe renal or hepatic disease, symptomatic heart disease, autoimmune disorders requiring immunosuppressive therapy, active thromboembolism or coagulopathy, or severe mental health condition (e.g., uncontrolled schizophrenia or depression with suicidal ideation), or other conditions at the discretion of the investigators.
6. Plans to move out of the study area within the next two years.

#### Eligibility considerations and justifications

Participants will be eligible beginning at age 16 years, as adolescent pregnancies are common in Guatemala and confer added stunting and NCD risk.^21^ Eligibility extends to 28 weeks gestation because most pregnant women in the study area first engage with prenatal care in the second trimester. Women with diabetes, serious medical or psychiatric conditions, or high-risk obstetric conditions will be excluded because these conditions may require specialized nutritional management beyond the scope of the counseling intervention designed for the general population of pregnant women. Women with known serious or lethal fetal anomalies (expected in <1% of pregnancies) will not be excluded, reflecting the trial’s pragmatic design and the elevated risk of subsequent maternal NCDs and mortality following pregnancy loss or stillbirth.^22^ Eligibility is not restricted by pre-pregnancy body mass index (BMI) because behavioral counseling is efficacious across BMI categories,^10^ and BMI-based eligibility criteria could be perceived as discriminatory and therefore were unacceptable to government stakeholders.

### Intervention

We will assess two evidence-based interventions during the pregnancy, postpartum, and early childhood periods: (1) food supplementation and (2) behavioral counseling to optimize maternal weight and promote healthy nutrition, physical activity, and infant feeding practices. The interventions are integrated as they will be delivered simultaneously and are theoretically complementary in achieving reductions in the household-level DBM. The combination is intended to capture the child growth benefits of supplementation while counteracting its potential to increase maternal gestational weight gain and postpartum weight retention.

Our conceptual model of the integrated intervention is based on the “capacity-load” model of NCDs (**Figure 1**).^23^ This model proposes that NCD risk increases when there is an imbalance between “metabolic capacity” and “metabolic load,” leading to dysregulated metabolic homeostasis. Metabolic capacity refers to physiologic traits developed in early life that are protective against NCDs. Metabolic load refers to behaviors and exposures that challenge metabolic homeostasis and lead to increased NCD risk. We hypothesize that our integrated intervention will reduce NCD risk among mothers and children through improved metabolic homeostasis: Food supplementation will increase metabolic capacity in infants by addressing factors that influence growth, including energy-inadequate diets and micronutrient deficiencies; behavioral counseling on healthy weight during pregnancy and postpartum will reduce metabolic load in mothers by addressing factors influencing weight gain, such as diet and physical activity.

**Figure 1:**
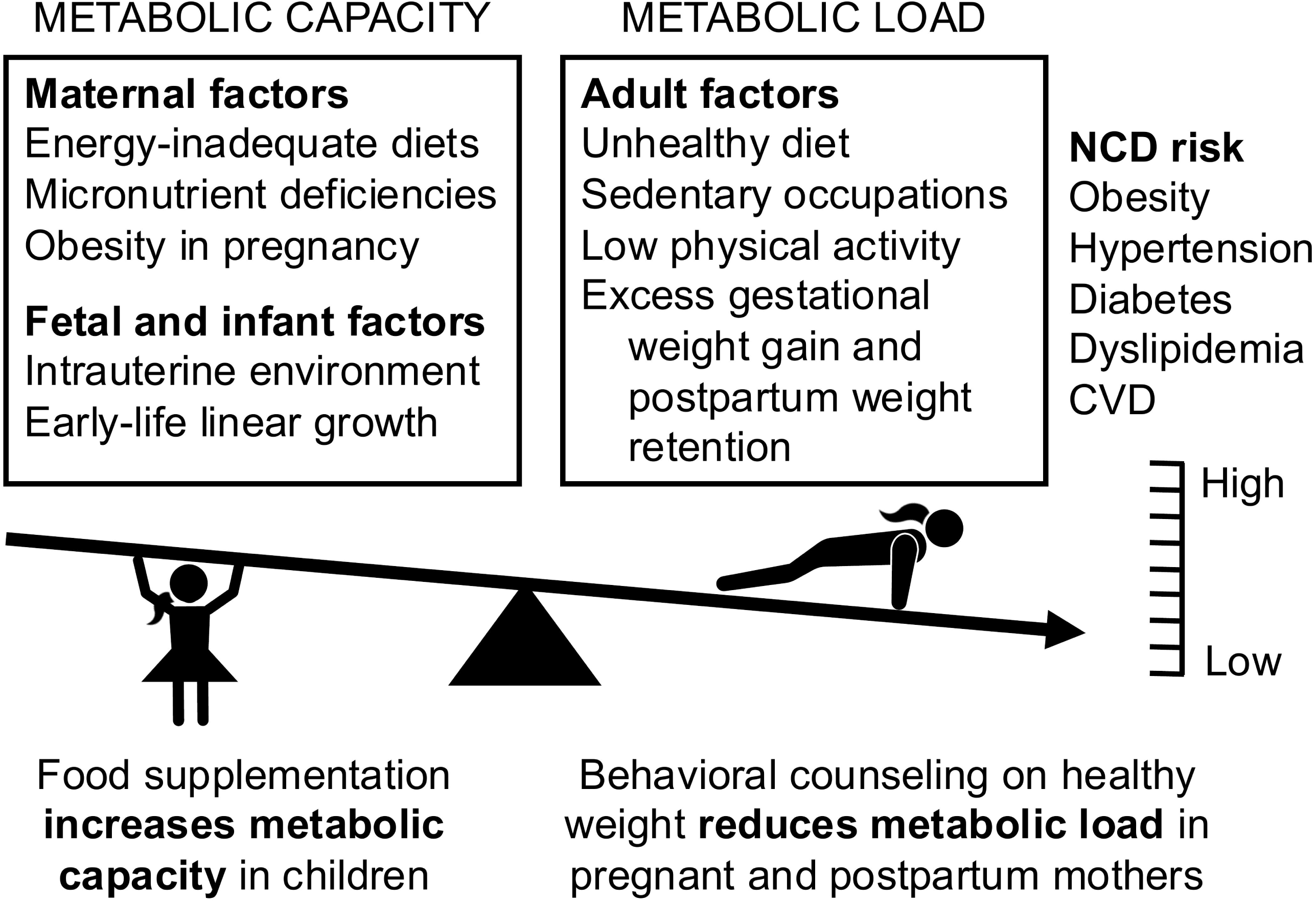
Conceptual model of the intervention based on the “capacity-load” model of non-communicable diseases

#### Intervention 1: Food supplementation

We designed the food supplement intervention to address challenges observed in our team’s prior research^24^ and modeling studies of optimal diets in rural Guatemala.^25^ In this setting, achieving adequate dietary quality is a greater challenge than meeting caloric or protein needs. Refined sugar consumption is high, and intake of fortified foods is limited. Food supplements are commonly shared within families. The composition of goods in the supplement was selected to prioritize locally available foods, low cost, and shelf life compatible with a monthly visit schedule, consistent with existing large-scale government programs. For example, the government’s healthy school meal program provides regular deliveries of a similar package including fresh food to over 3 million Guatemalan children.^26^

The food ration provides 148 kcal/day per capita, assuming sharing within a median family of 5 people (**Table 1**).^19,27^ The ration contains 5 food groups, the minimum dietary diversity requirement for a child aged 6-23 months or a reproductive-age woman.^28,29^ Macronutrient proportions (21% protein, 30% fat, 49% carbohydrate) align with WHO guidance for balanced supplementation in high-risk pregnancy.^30^ Eggs are included rather than meat due to cultural preferences, cost, and storage challenges.

**Table 1:** Food ration.

| Item | WHO food group<sup>28</sup> | Monthly ration | Energy/day per capita (kcal)<sup>d</sup> |
| --- | --- | --- | --- |
| Black beans | Pulses, nuts, and seeds | 5 lbs. | 51 |
| Chicken egg | Eggs | 45 eggs | 22 |
| Fortified blended flour | Grains, white roots, tubers, and plantains | 4 bags of 450g | 44 |
| Vegetable oil | N/A | 0.5 liter | 27 |
| Fruits and vegetables<sup>a</sup> | (1) Vitamin-A rich<sup>b</sup> and (2) others<sup>c</sup> | 5 lbs. | 2 |
| **Energy/day per capita (kcal)<sup>d</sup>** |  |  | **148<sup>e</sup>** |

<sup>a</sup>Based on seasonality, shelf life, and local acceptability to include at least 1 “vitamin A-rich” option and 1 “other” option in each delivery. <sup>b</sup>Vitamin A-rich fruits: Güicoy sazón, carrot, sweet potato, ayote, papaya, mango, dark leafy greens (chipilín, hierba mora/quilete/macuy, broccoli). <sup>c</sup>Other vegetables: tomato, cucumber, green bean, radish, beet, chile, green pepper, güisquil, güicoyito, cauliflower, cabbage. Other fruits: orange, pineapple, apple, watermelon. <sup>d</sup>Assumes 5 people share the ration and 30.4 days/month. <sup>e</sup>Value does not equal column sum due to rounding errors.

#### Intervention 2: Behavioral counseling

The behavioral counseling intervention was developed for the K’ASLEM trial and draws on Maya Health Alliance’s prior programs among Indigenous Maya populations in Guatemala, including maternal complementary and responsive feeding education^31,32^ and the promotion of healthy nutrition among adults with chronic diseases such as diabetes.^33^ Counseling consists of culturally and linguistically tailored home visits by trained educators, delivered monthly from enrollment through 12 months postpartum. Each visit lasts about one hour and is offered in Spanish or Kaqchikel, according to participant preference.

The behavioral counseling sessions integrate the Capability, Opportunity, Motivation, and Behavior (COM-B) model^34^ with Motivational Interviewing (MI),^35^ which is endorsed by the American College of Obstetricians and Gynecologists.^36^ Each visit includes (1) brief assessments of overall maternal and child health, including weight trajectory, diet, physical activity, and, during postpartum visits, infant feeding and growth; (2) review of goals from the prior visit; (3) identification of the primary session topic, tailored to each woman’s pregnancy or postpartum stage and specific needs; (4) delivery of educational content using a goal-focused activity; and (5) co-development of practical goals, using MI, for the participant to pursue after the session. The educational curriculum covers critical windows across the life course, spanning pregnancy, the early postpartum period (0-6 months), and the late postpartum period (6-12 months) (**Table 2**). Pregnancy topics include nutrition and dietary diversity, hydration and ultra-processed food consumption, physical activity, and birth preparedness. Early postpartum topics include postpartum self-care, exclusive breastfeeding, and preparation for complementary feeding. Late postpartum topics include safe complementary feeding practices, age-appropriate early stimulation, active play for mother and infant, dietary diversity reinforcement, birth spacing, and sustainability of healthy habits. Sessions are assigned based on identified need rather than in a fixed sequence and may be revisited if a previously addressed need recurs.

**Table 2:**
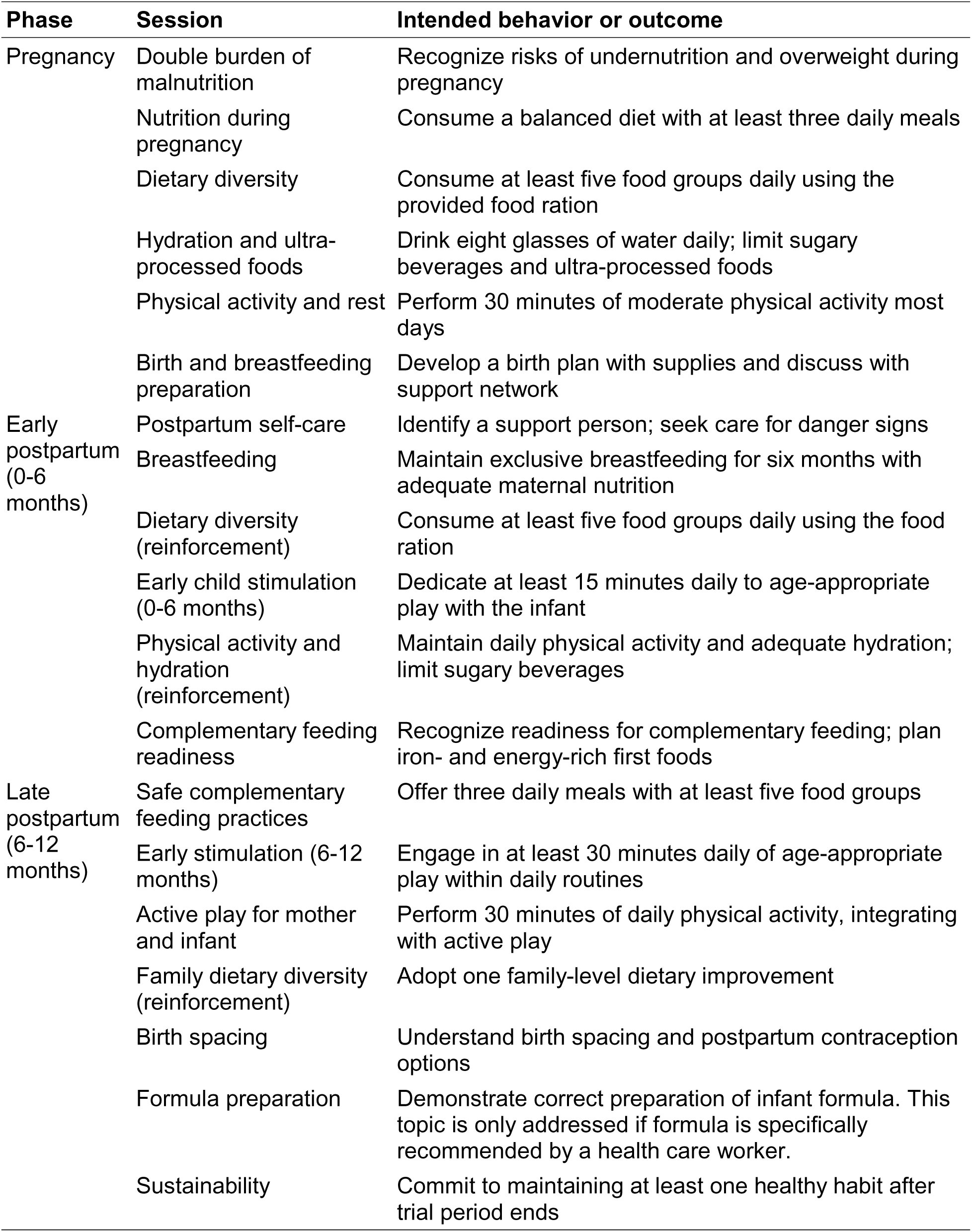
Educational topics and learning objectives.

Maternal weight counseling will be individualized. Gestational weight gain goals will be based on each participant’s BMI at enrollment as recommended by guidelines in Guatemala and the Institute of Medicine (**Table 3**);^37-39^ these goals will not be modified for women of short stature. After birth, mothers generally will be counseled to achieve a BMI within the normal range by 12 months postpartum. For women with overweight or obesity, an individualized weight target may be recommended.

**Table 3:** Gestational weight gain recommendations.

| **BMI category** | **Total weight gain in kg** | **Mean (range) weight gain in kg/week in 2nd and 3rd trimester** |
| --- | --- | --- |
| Underweight | 12.5-18 | 0.51 (0.44-0.58) |
| Normal | 11.5-16 | 0.42 (0.35-0.50) |
| Overweight | 7-11.5 | 0.28 (0.23-0.33) |
| Obesity | 5-9 | 0.22 (0.17-0.27) |

Individuals recruited as educators will be bilingual in Spanish and Kaqchikel Mayan. Educator training will be based on pedagogical approaches developed and validated in our prior trials.^31,40^ Initial training will last two weeks and consist of morning didactics and afternoon interactive activities. Competence will be assessed by post-training evaluation. We will conduct random field audits monthly for each educator and hold monthly one-day team refresher workshops, auditing, and supervision of educators.

### Comparator

The comparator will be enhanced usual care, which also will be available to intervention arm participants.

#### Usual care

Usual care is defined as the care provided by the Ministry of Health at no cost. Such care includes routine prenatal and postpartum care, routine infant preventive care, and hospital care for mothers and infants. National policy also recommends 2 kg monthly deliveries of fortified blended flour to children 6 to 12 months in municipalities where the trial will conduct activities.^16^

##### Enhancements to usual care

There are two enhancements. First, participants in both arms will be enrolled in Maya Health Alliance’s care navigation program. This program supports women and infants with high-risk or emergency conditions during pregnancy, birth, and the postpartum period.^41^ For emergencies, the program activates emergency transportation, accompaniment, and care coordination. For non-urgent high-risk conditions, navigators provide personalized referrals to the nearest government health center or hospital. Second, the study team will ensure that all children in the comparator group receive a 2 kg monthly delivery of fortified blended flour from 6 to 12 months of age. This enhancement addresses the possibility of stockouts or gaps in government supply that could otherwise leave comparator group children without access to this intervention. Children in the intervention arm already receive fortified blended flour as part of the food ration and therefore do not require additional provision.

### Outcomes

#### Co-primary outcomes

The co-primary outcomes are:

1. Maternal weight (kg) at 12 months postpartum (mean);
2. Child length-for-age z-score (LAZ) at 12 months of age using the WHO growth standard^42^ (mean).

#### Secondary outcomes

The secondary outcomes are:

1. Child stunting (LAZ <-2)^42^ at 12 months (proportion);
2. Child overall global development score from the Caregiver Reported Early Development Instruments (CREDI) long form raw scaled score^43^ at 12 months of age (mean);
3. Maternal overweight or obesity (BMI ≥25 kg/m^2^) at 12 months postpartum (proportion);
4. Maternal hemoglobin (g/dL) at 12 months postpartum (mean).

#### Exploratory outcomes

A full list of exploratory outcomes and their definitions are included in the Statistical Analysis Plan. Exploratory maternal outcomes include all-cause mortality, gestational weight gain,^39^ physical activity based on accelerometry, dietary diversity,^29^ food insecurity,^44^ health-related quality of life,^45^ blood pressure, anemia,^46^ hemoglobin A1c, fasting glucose, and fasting lipids. Exploratory child outcomes include stillbirth, infant mortality and morbidity, estimated birth weight,^47^ weight-for-age, weight-for-length, head circumference, breastfeeding indicators,^28^ and child feeding indicators.^28^

### Sample size

The planned sample size is 766 women in the 2 arms (n=383 per arm). We allow for up to 20% attrition based on our prior trials.^31,32,40^ Our assumed attrition includes women with another pregnancy by 12 months postpartum who will be excluded from the maternal co-primary outcome (estimated to be 5% of participants). Women with a pregnancy loss or infant death will be invited to continue, as they have a high risk of subsequent NCDs and mortality. Studies of similar maternal postpartum weight loss interventions in low-income, predominantly Latino U.S. populations found differences of 1.6-2.3 kg (standard deviation [SD] 5-11) at 12 months.^48,49^ A total of 318 women in each arm after attrition (80% of 383) gives us 80% power to detect a conservative yet clinically meaningful 1 kg (SD 4.5) mean difference between arms.

The planned sample size allows us to detect a mean difference of 0.25 (SD 1.0) length-for-age z-score (LAZ) at 12 months with 80% power. This effect size is in the range of other trials providing supplementation during pregnancy and early childhood.^50-53^

### Recruitment and retention

#### Recruitment procedures

The trial will use a rolling recruitment strategy to enroll approximately 43 participants per month over 18 months, for a total of 766 pregnant women and 766 infants. We anticipate that pregnant women will be recruited mainly through referrals from Ministry of Health facilities, traditional Indigenous midwives, and Maya Health Alliance’s clinical programs. Community engagement will support timely recruitment through these channels. At least one month before recruitment begins in each community, the study team will present the project to key authorities, including municipal officials, Community Development Councils (Spanish: “Consejos Comunitarios de Desarrollo” [COCODES]), and Ministry of Health leaders. Approval from these authorities will be required before initiating activities in any community.

#### Retention procedures

To maximize retention, we will collect multiple telephone contact numbers, record detailed information on the location of each participant’s home, and offer considerable flexibility regarding visit scheduling. If a woman becomes pregnant again during the study follow-up period, she will be excluded from the co-primary maternal outcome of weight at 12 months postpartum. However, she will be invited to continue participating in the trial and will continue to receive food supplementation and educator visits tailored to her current pregnancy stage and need. The infant from the index pregnancy will continue to be followed and assessed for the co-primary child outcome of length-for-age at 12 months of age. In the event that a woman experiences a pregnancy loss or infant death after enrollment, she will be invited to continue participating until 12 months after the delivery date or the date of fetal loss. If a maternal death occurs, the child will be followed until 12 months after birth, provided new consent is obtained. If a woman in the intervention group develops diabetes (gestational or non-gestational) during the study, dietary counseling will be discontinued; the woman will continue to receive home visits and the food ration, and she also will be referred to the local health facility for nutritional management. Women in the comparator group who develop diabetes also will be referred to the health center. In both cases, women will remain enrolled and continue study visits.

### Randomization and allocation

#### Randomization procedure

Following consent, pregnant women will be individually randomized to the intervention or comparator arm in a 1:1 allocation, using permuted blocks of 4, 6, or 8 participants. The study statistician will use Stata to generate the randomization table, which will be uploaded to REDCap’s randomization module. After a participant has completed enrollment and baseline assessments, the study nurse will apply the randomization function in REDCap to reveal the participant’s group assignment.

#### Blinding

The randomization sequence will be concealed in REDCap and revealed only at the moment of randomization; this concealment, together with variable block sizes, prevents recruitment nurses from anticipating assignments before enrollment. After allocation, the following study members will be unblinded: educators delivering the behavioral intervention, logistics staff delivering food rations, recruitment nurses, and study supervisors. The nurses who collect outcome data will be blinded. To preserve blinding of outcome assessors, participants will be instructed not to disclose their allocation during outcome visits, outcome nurses will maintain workflows independent of staff delivering the intervention, and no forms provided to participants will record group allocation. The study statistician will remain blinded until the primary analysis dataset is locked.

### Participant timeline

Study participants will have five scheduled study visits for data collection (**Figure 2**): (1) prior to 28 weeks gestation, (2) at 36 weeks gestation, (3) within 7 days after delivery, (4) at 6 months postpartum, and (5) at 12 months postpartum. The schedule of enrollment, allocation, interventions, and assessments is shown in **Table 4**.

**Figure 2:**
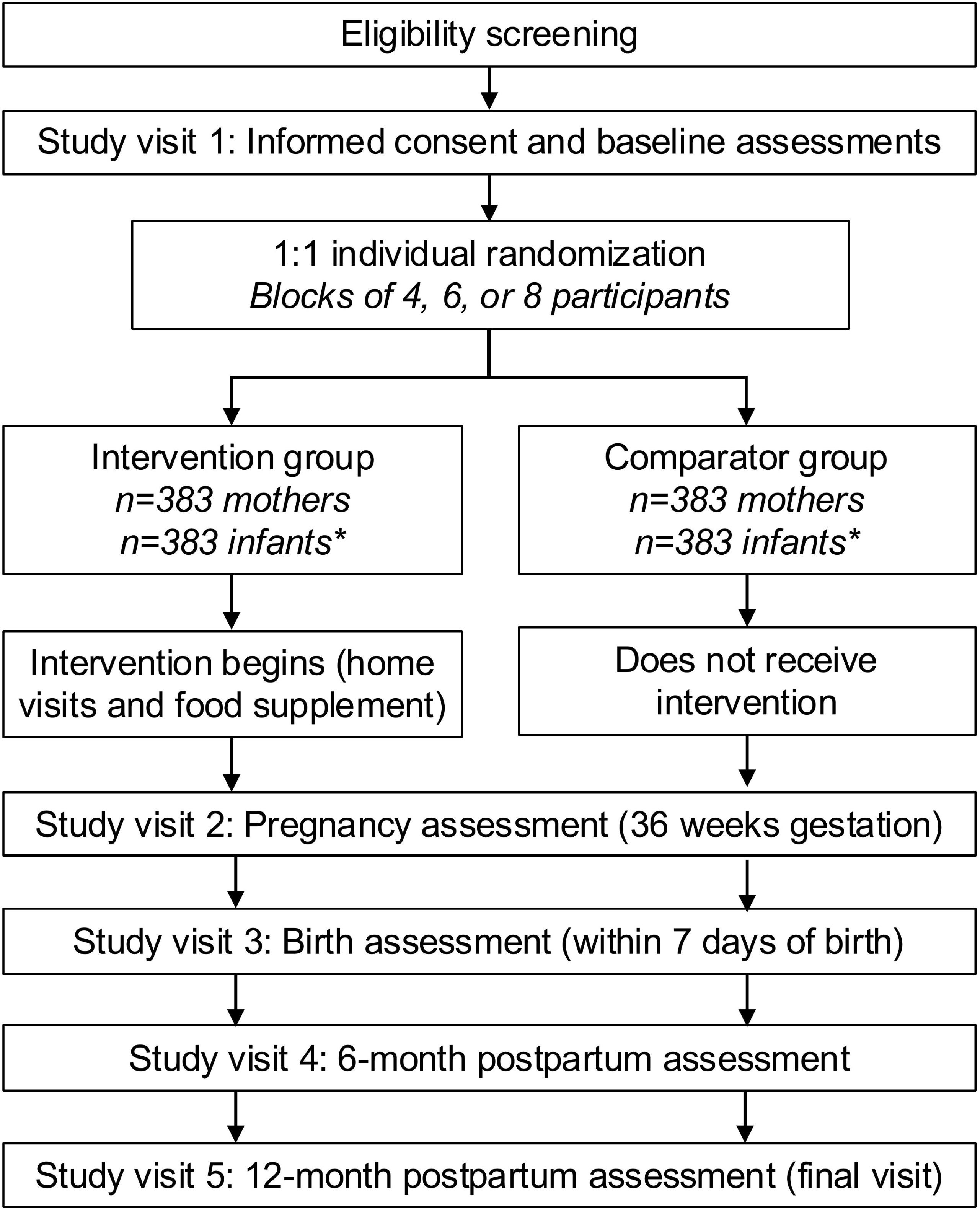
Schematic of participant timeline

**Table 4:**
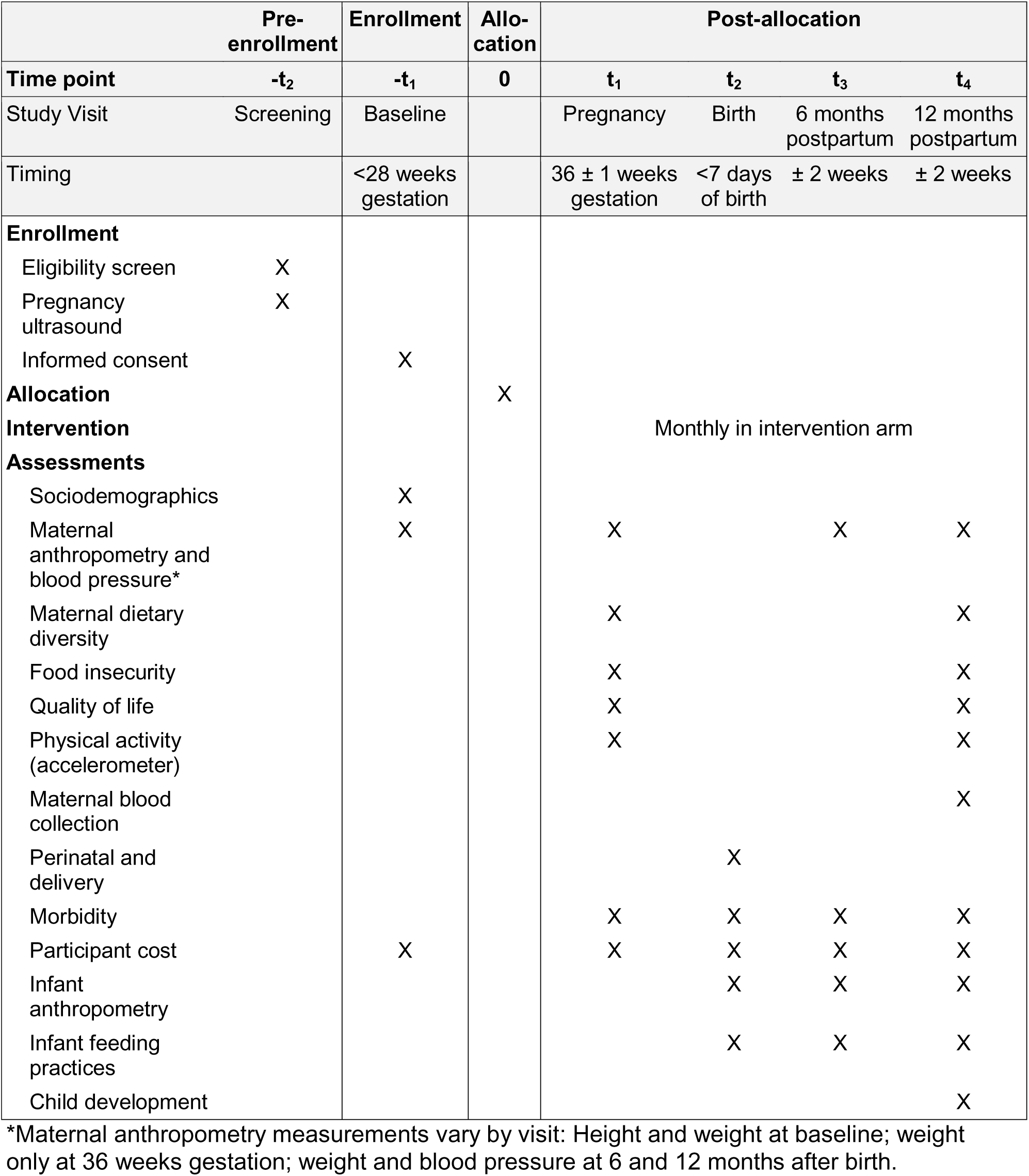
Schedule of enrollment, allocation, interventions, and assessments.

### Data collection

Outcome data will be collected by bilingual (Spanish-Kaqchikel) study nurses trained in questionnaire administration, anthropometry, and venous blood collection. Study visits will occur at different times than educator visits. Study visits will generally be conducted in participants’ homes, though visits may also be conducted in alternative private locations such as community centers, churches, or other suitable spaces per participant preference. In cases where childcare responsibilities are shared, we will ask the mother if she would like to invite other caregivers to be present during study visits to assist in answering questions on topics such as feeding practices, child development, or health care utilization.

Most data, including structured questionnaire responses, will be entered directly into REDCap^54^ in the field using tablets. Questionnaire data will include self-reported information described in the outcomes section, as well as baseline assessments of maternal sociodemographic characteristics, household socioeconomic status,^55^ and medical and obstetric history. Baseline questionnaires were selected to enable comparisons with prior surveys and nutrition trials conducted in Guatemala.^20,56,57^

Anthropometric measurements will be taken in triplicate by study nurses trained and standardized following INCAP guidelines, and standardization exercises will be repeated at least every 4 months to maintain measurement reliability. Maternal height will be measured using a portable stadiometer and maternal weight using a digital scale placed on a firm, level surface and confirmed to read zero before each measurement session. Infant length will be measured using a portable length board, weight using a hanging scale (Seca 310, Seca) zeroed with the empty sling before each session, and head circumference using a nonelastic measuring tape. Maternal blood pressure will be measured based on a standardized procedure using a validated digital monitor with appropriate cuff size (HEM907XL, Omron) and positioning. Physical activity will be assessed by accelerometry. Participants will wear the ActiGraph LEAP watch on their wrist during waking hours over a seven-day period, except during activities involving full-body water contact. At 12 months postpartum, maternal venous blood will be collected after at least 8 hours of fasting by antecubital venipuncture using standard aseptic technique. Samples will be transported at room temperature to the processing laboratory within 60 minutes of collection, centrifuged within 4 hours, and stored at -20°C before transport via cold chain to INCAP’s laboratory for analysis on the Roche Cobas C111 system. The study coordinator will conduct weekly quality control checks on 10% of study visits and random field audits.

### Statistical methods

A detailed Statistical Analysis Plan accompanies this protocol. Statistical methods are guided by the estimand framework.^58^ For each primary and secondary outcome, the Statistical Analysis Plan defines the target population, handling of intercurrent events (events after randomization that complicate interpretation, such as a subsequent pregnancy or a death),^59^ statistical methods, handling of missing data, and prespecified subgroup and sensitivity analyses. The primary analyses will compare the two trial arms on the co-primary outcomes at 12 months using regression models with baseline covariate adjustment, reporting adjusted mean differences with 95% confidence intervals. Child length-for-age z-score will be analyzed in all randomized children following the intention-to-treat principle. Maternal weight will be analyzed in a modified intention-to-treat population that excludes women with a subsequent pregnancy before the 12-month assessment. Each co-primary outcome will be tested independently at the full alpha (α = 0.05, two-sided) with no multiplicity correction; the trial will be considered positive only if both co-primary outcomes reach statistical significance.^60^ Secondary outcomes will be analyzed using regression models consistent with the primary analysis framework. No interim efficacy analyses are planned. Missing data will be handled primarily with multiple imputation.

### Implementation research plan

Even if the intervention evaluated in the trial were effective, its public health impact would be limited without understanding the factors that shape widespread implementation. Our implementation research plan aims to identify these factors and develop a package of strategies for evaluation in a future large-scale implementation study (e.g., a Hybrid Type 2, Hybrid Type 3, or implementation trial). The plan is guided by Implementation Mapping, a systematic methodology for developing strategies to enhance intervention adoption, implementation, and sustainability.^61^ We will conduct Steps 1 through 3; the remaining steps will be reserved for the future study (**Figure 3**).

**Figure 3:**
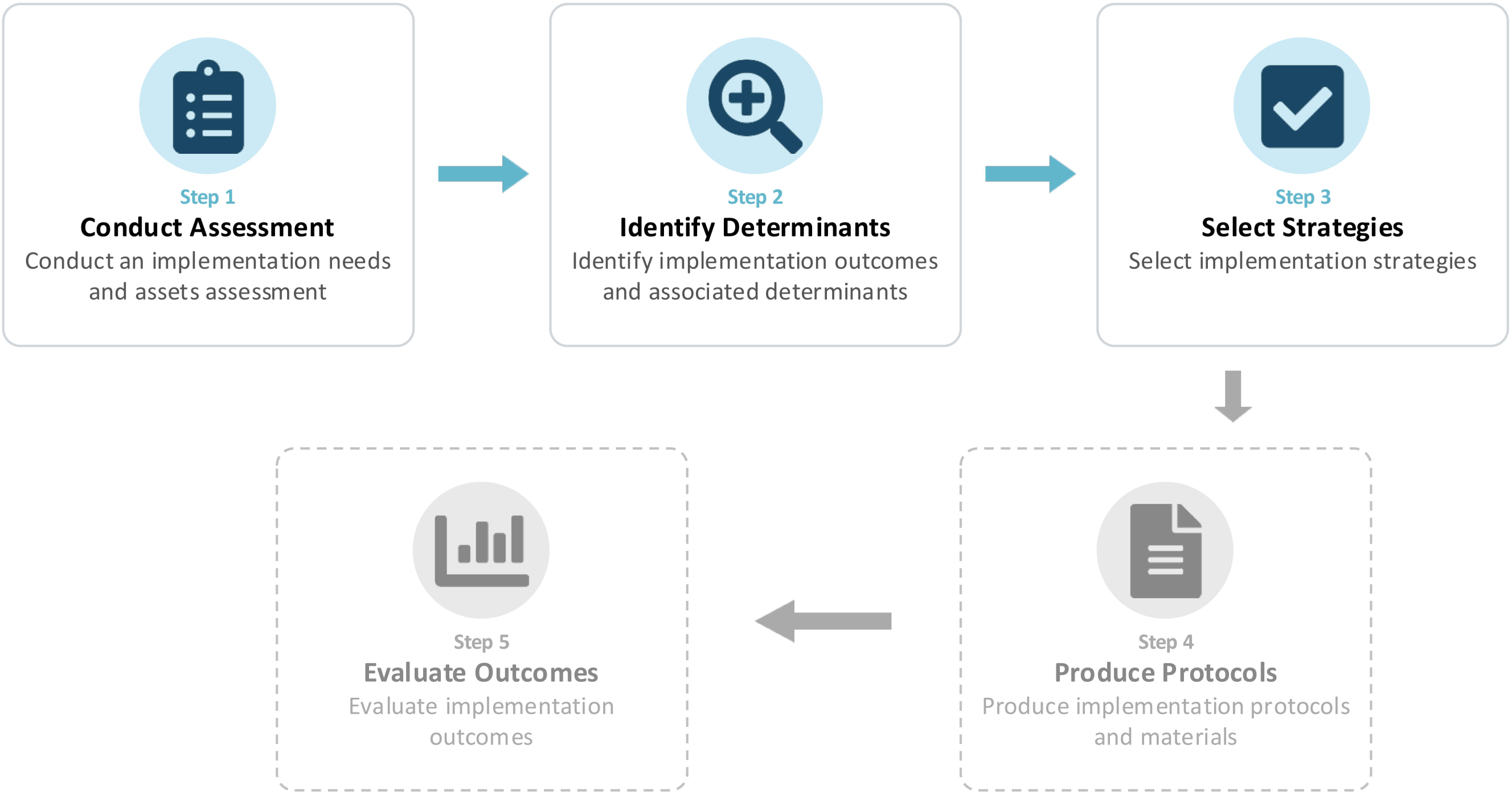
Implementation mapping steps

Step 1 will be a needs and assets assessment to identify determinants of implementing the intervention at scale. Data sources will include quantitative implementation outcomes guided by RE-AIM (**Table 5**),^62^ weekly study team meeting recordings, educator field notes, focus groups with trial staff, and semi-structured interviews with key informants. Findings from the data sources will be integrated and analyzed using the Tailored Implementation for Chronic Diseases (TICD) checklist.^63^ In Step 2, we will define performance objectives that would be most important in the future large-scale study. A performance objective is a concrete action that a specific actor would need to take for a given determinant to be addressed. In Step 3, we will convene a multidisciplinary working group to select implementation strategies matched to the performance objectives and implementation outcomes. This process will be guided by the Implementation Research Logic Model.^64^ For the highest-rated strategies, the working group will specify operational details using the strategy specification framework outlined by Proctor et al.^65^ Upon completion of these steps, we will have produced the foundational materials needed to design the future large-scale implementation study.

**Table 5:** Implementation outcomes guided by RE-AIM.

| **RE-AIM domain** | **What is assessed** | **Data sources** |
| --- | --- | --- |
| Reach | Proportion of eligible women approached who choose to participate | Trial enrollment data |
|  | Representativeness of participants vs. (a) general population and (b) prior nutrition trials in Guatemala | Trial enrollment data, health surveys, census microdata, data from prior trials |
| Effectiveness | See primary and secondary outcomes | Outcome assessments |
| Adoption | Proportion of health facilities referring at least 1 participant for trial enrollment | Trial enrollment data |
| Implementation | Fidelity: Proportion of scheduled visits delivered | Educator visit logs |
|  | Fidelity: Degree to which the intervention is delivered as intended | Scoring of audio recordings of educator sessions |
|  | Contact intensity: Total number of contact hours | Educator visit logs |
| Maintenance | Costs and cost-effectiveness | Economic evaluation |
|  | Participant dropout rate | Trial enrollment data |
|  | Characteristics of completers vs. non-completers | Trial enrollment data |

### Economic evaluation

Our economic evaluation will provide information to policymakers for implementation and budgeting. Consistent with the trial’s focus on effectiveness (Hybrid Type 1), we will conduct a within-trial cost-effectiveness analysis comparing the intervention to enhanced usual care. The time horizon will be from enrollment in the second trimester to 12 months postpartum. We will estimate costs for usual care, enhancements to usual care, and the intervention using costing tools previously developed by our team in Guatemala.^40^ For each, we will estimate component costs (system-level, patient and family, and health worker), total costs, and cost per beneficiary, disaggregating system-level costs across the six WHO health system building blocks. All costs will be valued at local market rates in constant 2026 US dollars. Incremental costs will be estimated with generalized linear models appropriate for skewed cost data, with cost per beneficiary as the outcome and study arm as the exposure; 95% confidence intervals will be obtained by nonparametric bootstrapping. Costs will be discounted at 3% (0-6% in sensitivity analyses). Effectiveness will use the co-primary continuous outcomes (maternal weight, child length-for-age z-score) and dichotomized measures (maternal overweight/obesity, child stunting), with incremental effectiveness from linear or logistic regression. We will report incremental cost-effectiveness ratios and cost-effectiveness acceptability curves across willingness-to-pay thresholds, from both the health system and societal perspectives.^66^

### Patient and public involvement

Patients and the public were not involved in the development of the protocol. Their involvement in the dissemination of findings is detailed in the Dissemination plan.

## ETHICS AND DISSEMINATION

### Research ethics approval

Ethics approval was obtained from the Maya Health Alliance IRB (August 2025), the INCAP Research Ethics Committee (MI-CIE-26-007), and the Mass General Brigham IRB (2024P003339), which serves as the single IRB of record for U.S. collaborating institutions.

### Informed consent

Trained bilingual field staff will obtain verbal informed consent in the participant’s preferred language. Verbal consent is culturally appropriate because many Indigenous Maya people in Guatemala have a deeply rooted distrust of signing documents. Staff will read a consent script aloud, assess comprehension using a teach-back process before proceeding, and document consent by signing and dating the form; participants receive a printed copy. Separate verbal consent will be obtained before blood collection. For mothers under 18 years of age, a parent or legal guardian confirmed using a national identity document number will provide written consent, and the adolescent will provide verbal assent. In the event of a maternal death, study staff will ask the child’s legal guardian whether they wish to provide renewed consent for the child’s continued involvement in the study; this discussion will begin only after a period of bereavement and with sensitivity to the family’s cultural and grieving practices.

### Data monitoring and safety

The trial poses not more than minimal risk as defined in U.S. federal research regulations (45 CFR 46.102), and no serious adverse events attributable to trial procedures are anticipated. Nevertheless, pregnant Indigenous women and infants in rural Guatemala are vulnerable populations who require special protections. An independent Data and Safety Monitoring Board (DSMB) with multidisciplinary expertise was established to provide oversight of participant safety, trial conduct, and data integrity. The investigator team will track and report to the IRBs, DSMB, and funding agency any health complications, perceived adverse events, or complaints reported by participants or community members. Fatal or life-threatening serious adverse events will be reported to the IRBs and DSMB within 3 days and other serious or unanticipated events within 7 days. Stillbirth, infant death, and maternal death occur at appreciable background rates in this population and will be recorded as serious adverse events irrespective of attribution, reviewed by the DSMB, and reported by arm.

### Dissemination plan

Results will be published in peer-reviewed, open-access journals following established reporting guidelines and will be presented at national and international conferences. De-identified datasets, study instruments, and training materials will be deposited in a public data-sharing repository by the time of primary publication. In Guatemala, findings will be presented to the Ministry of Health and the Secretariat of Food and Nutritional Security. Across Central America, findings will also be shared through public-sector technical networks in which INCAP participates. The study team will return plain-language summaries in Spanish and Kaqchikel to participants and hold public dissemination meetings with community leaders at each site to discuss the findings and their implications.

## ADDITIONAL INFORMATION

### Authors’ contributions

DF and MMazariegos are co-first authors and jointly drafted the trial protocol. MR-Z and PR are co-senior authors, the project’s co-principal investigators, and jointly supervise the trial’s conduct. PR, MR-Z, DF, CL, MMazariegos, ACM, and MMoniz contributed to study design and protocol content in their areas of expertise. YJM, SHH, and WARG contributed to protocol refinement, intervention adaptation, and the overall development of trial procedures. All authors reviewed and approved the final manuscript.

## Supporting information

SPIRIT checklist

Strobe checklist

Statistical Analysis Plan

## Data Availability

Data will be made available online after the trial concludes

