## Supplementary material for "Food supplementation and behavioral counseling to reduce the double burden of malnutrition among pregnant women and their infants: protocol for the K’ASLEM hybrid type 1 effectiveness-implementation randomized controlled trial": Statistical Analysis Plan

An integrated intervention to address the double burden of malnutrition in Guatemala (K’ASLEM trial)

**Study Acronym / Short Title:** K’ASLEM

**SAP Version:** 1.0

**SAP Date:** 29 April 2026

**Protocol Version Referenced:** Version 3.0, March 17, 2025

**IRB/Ethics Approval Number:** Mass General Brigham (2024P003339), INCAP (CIE-REV 141/2025), Maya Health Alliance (August 2025)

**ClinicalTrials.gov Identifier:** NCT06750120

Signature Block

| **Role** | **Name** | **Signature** | **Date** |
| --- | --- | --- | --- |
| Principal Investigator (contact) | Peter Rohloff |  |  |
| Principal Investigator | Manuel Ramirez-Zea |  |  |
| Principal Investigator | David Flood |  |  |
| Lead Biostatistician | Ann C Miller |  |  |
| Co-Investigator | Mónica Mazariegos |  |  |

**DOCUMENT VERSION HISTORY**

| **Version** | **Date** | **Author** | **Description of Changes** |
| --- | --- | --- | --- |
| 1.0 | [ ] | Ann C. Miller | Initial SAP — pre-specified prior to database lock |

This SAP was finalized BEFORE unblinding. Any post-hoc amendments must be documented with full justification

### 1. Introduction and Study Background

#### 1.1. Background and Rationale

Adult populations globally are experiencing rapid increases in non-communicable diseases (NCDs) attributable to overnutrition, yet child undernutrition persists at extraordinary levels. This “double burden of malnutrition” (DBM) is commonly defined as maternal overweight/obesity (body mass index ≥25 kg/m^2^) and child linear growth failure (“stunting,” or length-for-age z-score <-2) in a mother-child dyad. This form of DBM occurs in more than 20% of households in some low- and middle-income countries (LMICs) and is disproportionately concentrated among socioeconomically disadvantaged households.^1-3^ Pregnancy, postpartum, and early childhood are critical life stages in which to address the DBM. Among children, linear growth failure during these stages is associated with profound biological, developmental, and economic consequences throughout the lifespan, including an increased risk of NCDs.^4-7^ Among mothers, excess gestational weight gain and postpartum weight retention increase risk of pregnancy complications, NCDs, and mortality.^8^

Evidence-based interventions exist that prevent child growth failure or promote optimal maternal weight, but critical knowledge gaps remain in implementing them as integrated, sustainable, and accessible solutions among the world’s poorest.^9^ Two interventions with strong evidence are (1) food supplementation during pregnancy and early postpartum for mothers, and for both mothers and infants after 6 months upon the transition to complementary foods;^10,11^ and (2) behavioral counseling to optimize gestational weight gain and limit postpartum weight retention.^12-15^

Despite the proven efficacy of these interventions in isolation, their population benefits have been limited for a few key reasons. Food supplementation improves child growth but may cause excessive maternal weight gain during and after pregnancy.^9,16^ Implementation knowledge gaps also have limited the scale-up of effective nutrition interventions in LMICs. Efficacy trials of nutrition supplementation have generally paid limited attention to implementation or sustainability,^17^ and behavioral counseling trials among pregnant or postpartum women have been conducted almost exclusively in high-income countries.^18^ Prior trials have often excluded the populations most affected by the DBM through design choices, which limit generalizability among the most at-risk households.^19^

#### 1.2. Study Objectives

##### 1.2.1. Primary Objective

To evaluate the effect of a combined behavioral education and food supplementation intervention — initiated during pregnancy and continuing through 12 months postpartum — on co-primary endpoints in mother-child dyads, compared to enhanced usual care: (1) child length-for-age z-score (LAZ) at 12 months of age, and (2) maternal weight in kg 12 months postpartum.

##### 1.2.2. Secondary Objectives

- To evaluate the effect of the intervention on secondary maternal and infant outcomes.
- To assess maternal and child exploratory outcomes.
- To evaluate the effect of the intervention in prespecified subgroups

#### 1.3. Purpose of this Document

This Statistical Analysis Plan (SAP) details all pre-specified analyses to be conducted for the primary, secondary, and exploratory objectives of this trial. It is intended to be read in conjunction with the study protocol (Version [.1], dated [April 23, 2026]). In the event of any discrepancy, the SAP takes precedence for analytical decisions. This document was finalized prior to database lock and unblinding.

### 2. Study Design Overview

| **Design Element** | **Specification** |
| --- | --- |
| Study Design | Parallel-arm, randomized controlled trial |
| Unit of Randomization | Pregnant woman-child dyad |
| Arms | Intervention (behavioral education + nutritional supplementation) vs. comparator (enhanced usual care) |
| Allocation Ratio | 1:1 |
| Randomization Method | Block randomization, in randomly allocated blocks of 4, 6 or 8. |
| Masking | Open-label for participants, educators, and supervisors. Blinded:  (1) study nurses conducting outcome data collection  (2) study statistician and data manager, until primary analysis dataset is locked |
| Primary Follow-up | 12 months post-delivery |
| Assessment Timepoints | Enrollment (prior to 28 weeks gestation), 36 weeks gestation, within 7 days after delivery, 6 months postpartum, 12 months postpartum |
| Target Sample Size | 766 dyads (383 per arm) |
| Target Population | Pregnant women (up to 28 weeks gestational age) and their infants, followed for 12 months after delivery |

In our protocol, we have two co-primary hypotheses, and 4 secondary hypotheses. Our main efficacy endpoints are to assess the coprimary outcome of Child length-for-age z score (LAZ) at 12 months of age using WHO growth standard AND maternal weight in kilos at 12 months postpartum by study arm. Each of the two co-primary endpoints will be tested independently at the full alpha (α = 0.05, two-sided).

| **ID** | **Outcome** | **Sample** | **12 month endpoint (Main efficacy)*** |
| --- | --- | --- | --- |
| 1 | Co-Primary | Full sample of children | H01: There is no difference in child LAZ score. |
| 2 | Co-Primary | Women who have not had a second pregnancy within 12 months after the study child’s birth | H02: There is no difference in mean maternal weight in kilos at 12 months post partum between study arms among women who have not had a second pregnancy. |
| 3 | Secondary | Only children | H03: There is no difference in proportion of child stunting (LAZ<-2) at 12 months by study arm |
| 4 | Secondary | Only children | H04: There is no difference in child development as measured by CREDI longform total scores at 12 months by study arm. |
| 5 | Secondary | Only women | H05: There is no difference in the proportion of women with BMI >=25 kg/m^2) at 12 months by study arm. |
| 6 | Secondary | Only women | H06: there is no difference in mean maternal hemoglobin (g/dL) at 12 months by study arm. |

### 3. Endpoints and Estimands

#### 3.1. Co-Primary Endpoints

These endpoints are designated co-primary under the logical AND framework: the intervention will be declared successful only if BOTH endpoints reach statistical significance at the pre-specified alpha level. Each endpoint is tested independently at the full alpha (α = 0.05, two-sided). This framework controls the familywise Type I error rate; no multiplicity correction between the two co-primary endpoints is applied.

| **Endpoint** | **Population** | **Timepoint** |
| --- | --- | --- |
| CP-1: Infant length-for-age z-score | Index child of enrolled dyad | 12 months post delivery |
| CP-2: Maternal weight in kilograms | Pregnant woman enrolled in the trial at <28 weeks gestation | 12 months post delivery |

**Success criterion:** The intervention is considered effective if and only if BOTH CP-1 AND CP-2 achieve p < 0.05 (two-sided) in their respective primary analyses. A significant result on only one co-primary endpoint does not constitute trial success. We will first present a comparison of proportions of subsequent pregnancies prior to the 12 month postpartum follow up per study arm. We will exclude women who have second pregnancies from the maternal endpoint analysis, using a mITT design.

#### 3.2. Secondary Endpoints

| **ID** | **Endpoint Description** | **Dyad Member** | **Timepoint** |
| --- | --- | --- | --- |
| S-1 | Infant stunting (LAZ<-2) proportion at 12 mo | Child | 12 months of age |
| S-2 | Infant global development score on CREDI long form at 12 months (mean) | Child | 12 months of age |
| S-3 | Maternal BMI >=25 kg/m^2 proportion | Mother | 12 months postpartum |
| S-4 | Maternal hemoglobin (g/dL) at 12 months postpartum (mean) | Mother | 12 months postpartum |

Secondary endpoints are considered supportive and exploratory. No formal multiplicity adjustment is applied, but results will be interpreted in the context of the number of tests performed. p-values and confidence intervals will be reported for all secondary analyses.

#### 3.3. Exploratory Endpoints

##### 3.3.1. *Maternal exploratory outcomes:*

- All-cause maternal death from enrollment through 12 months postpartum (proportion)
- Maternal morbidity, defined as a woman experiencing one or more non-fatal serious adverse events from enrollment through 12 months postpartum (proportion)
- Adequate gestational weight gain velocity between enrollment and 36 weeks gestation, as defined by Institute of Medicine category^20,21^ (proportion)
- BMI (kg/m^2^) at 12 months postpartum (mean)
- Moderate-to-vigorous physical activity (minutes/day) by 7-day accelerometry at 36 weeks gestation and 12 months postpartum (mean)
- Minimum dietary diversity for women at 36 weeks gestation and 12 months postpartum^22^ (proportion)
- Food insecurity (Food Insecurity Experience Scale, 30-day household-level version^23,24^) at 36 weeks gestation and 12 months postpartum (mean score)
- Maternal anemia using WHO altitude-adjusted thresholds^25^ at 12 months postpartum (proportion)
- Health-related quality of life (PROMIS Global Health) at 36 weeks gestation and 12 months postpartum: Global Physical Health T-score (mean) and Global Mental Health T-score (mean)
- Systolic blood pressure (mmHg) at 12 months postpartum (mean)
- Diastolic blood pressure (mmHg) at 12 months postpartum (mean)
- Hypertension (systolic blood pressure ≥130 mmHg or diastolic blood pressure ≥80 mmHg)^26^ at 12 months postpartum (proportion)
- Maternal hemoglobin A1c (%) at 12 months postpartum (mean)
- Fasting plasma glucose (mg/dL) at 12 months postpartum (mean)
- Maternal diabetes (HbA1c ≥6.5% or fasting plasma glucose ≥126 mg/dL) at 12 months postpartum (proportion)
- Fasting lipid panel (total cholesterol, triglycerides, LDL, HDL, and VLDL [mg/dL]) at 12 months postpartum (mean)

##### 3.3.2. *Child exploratory outcomes:*

- Stillbirth, defined as fetal death at ≥28 weeks gestation^27^ (proportion of all births)
- Infant mortality, defined as death among live births within 12 months of birth (proportion)
- Infant morbidity, defined as an infant experiencing one or more non-fatal serious adverse events from birth through 12 months of age (proportion)
- Birth weight (g) estimated from postnatal weights using the Reeds 2 model,^28^ which was previously validated in the study setting^29^ (mean)
- Low birth weight (<2500 g) estimated from postnatal weights using the Reeds 2 model^28^ (proportion)
- Child length-for-age z score (LAZ)^30^ at birth and 6 months of age (mean)
- Stunting (LAZ <-2)^30^ at 6 months of age and severe stunting (LAZ <-3)^30^ at 6 and 12 months of age (proportion)
- Child weight-for-age z score (WAZ)^30^ at 6 and 12 months of age (mean)
- Underweight (WAZ <-2)^30^ at 6 months and 12 months of age (proportion)
- Child weight-for-length z score (WLZ)^30^ at 6 and 12 months of age (mean)
- Moderate acute malnutrition (WLZ <-2 to ≥-3)^30^ and severe acute malnutrition (WLZ <-3) at 6 and 12 months of age (proportion)
- Head circumference-for-age z score (HCZ)^30^ at birth, 6 months, and 12 months of age (mean)
- Breastfeeding indicators:^31^ Early initiation of breastfeeding within 1 hour of birth, exclusively breastfed for the first two days after birth, exclusive breastfeeding at 6 months, and continued breastfeeding at 12 months of age (proportion)
- Child feeding indicators:^31^ Minimum dietary diversity, minimum meal frequency, and minimum acceptable diet at 6 and 12 months of age (proportion)

#### 3.4. Estimand Framework (ICH E9(R1))

The following intercurrent event strategies are pre-specified in accordance with the ICH E9(R1) addendum:

| **Intercurrent Event** | **CP-1 strategy** | **CP-2 strategy** | **Rationale** |
| --- | --- | --- | --- |
| Subsequent pregnancy before 12-month follow-up | Primary: Treatment policy SA-1: Principal stratum (subpopulation: children of women without subsequent pregnancy) | Primary: Principal stratum (subpopulation: women without subsequent pregnancy; mITT) SA-1: Not applicable (CP-2 primary applies the SA-1 exclusion) | For CP-1, the child's LAZ remains measurable regardless of subsequent maternal pregnancy and is retained under treatment policy. For CP-2, subsequent pregnancy fundamentally alters the maternal weight trajectory, so the estimand of interest is the effect within the subpopulation of women without subsequent pregnancy. |
| Intervention discontinuation, participant remains in study | Primary: Treatment policy SA-4: Per-protocol sensitivity analysis (subpopulation: per-protocol adherers, defined in Section 6) | Primary: Treatment policy SA-4: Per-protocol sensitivity analysis (subpopulation: per-protocol adherers, defined in Section 6) | Treatment policy preserves the randomized comparison. SA-4 is a supportive per-protocol sensitivity analysis intended to describe the treatment effect among adherers as  observed. |
| Stillbirth | Primary: N/A | Primary: Treatment policy-.. | Stillbirth is not an intercurrent event for CP-1 because the unit of analysis is a live-born child. For CP-2, maternal weight remains measurable regardless of pregnancy outcome, so the mother is retained under treatment policy. Stillbirth is reported by arm as an exploratory endpoint. |
| Child death (live-born)* | Primary: Principal stratum (subpopulation: children surviving to 12 months) Note: Post-birth, child’s baseline data may be used to inform estimates during multiple imputation | Primary: Treatment policy | LAZ is not biologically defined for a deceased child, so the CP-1 primary uses a principal stratum estimand defined on the subpopulation of children surviving to 12 months. This estimand is interpretable under the assumption that allocation does not differentially affect child survival. For CP-2, the bereaved mother's weight remains measurable, and bereavement effects on weight are retained as part of the randomized exposure under treatment policy. |
| Maternal death* | Primary: Treatment policy | Primary: Principal stratum (subpopulation: mothers surviving to 12 months postpartum) | For CP-1, the child's LAZ remains measurable on a surviving child even if the mother dies, so treatment policy applies and the estimand is the effect of randomized exposure on child LAZ regardless of maternal survival. For CP-2, maternal weight is not measurable for deceased mothers, so the principal stratum estimand is defined on the subpopulation of mothers surviving to 12 months postpartum and is interpretable under the assumption that allocation does not differentially affect maternal survival. |
| Loss to follow-up or withdrawal of consent | Primary: Treatment policy with multiple imputation under MAR SA-2: Complete case SA-3: MNAR | Primary: Treatment policy estimand; multiple imputation under MAR SA-2: Complete case SA-3: MNAR | See Section 8 (Missing Data) |

*The CP-1 estimand under child death and the CP-2 estimand under maternal death rely on the assumption that randomization does not differentially affect survival. Given the small expected number of deaths in this trial, this assumption is not likely to be testable. Mortality will be reported by arm as an exploratory endpoint.

### 4. Sample Size and Power

#### 4.1. Sample Size Calculation

##### 4.1.1. Infant Endpoint (CP-1)

| **Parameter** | **Value** |
| --- | --- |
| Effect size (expected difference) | 0.25 LAZ |
| Standard deviation (if continuous) | 1.0 |
| Test type | Multiple linear regression |
| Alpha (two-sided) | 0.05 |
| Power | 80% |
| Required dyads per arm | 318 |
| Planned sample size (inflated for expected attrition) | 383 |

##### 4.1.2. Maternal Endpoint (CP-2)

| **Parameter** | **Value** |
| --- | --- |
| Effect size (expected difference) | 1 kg |
| Standard deviation (if continuous) | 4.5 kg |
| Test type | Multiple linear regression |
| Alpha (two-sided) | 0.05 |
| Power | 80% |
| Required dyads per arm | 318 |
| Planned sample size (inflated for expected attrition) | 383 |

Of note, the planned sample size of 383 per arm was derived by inflating the 318 required dyads per arm by approximately 20% (318 × 1.20 ≈ 383) to account for expected attrition from both loss to follow-up and for exclusion of women with a subsequent pregnancy from the CP-2 primary analysis. The 318 required dyads per arm listed for CP-1 reflects the power constraint from CP-2, not an independent CP-1 calculation (CP-1 achieves approximately 89% power at this sample size).

### 5. Randomization and Allocation

#### 5.1. Randomization Procedure

Following consent, pregnant women will be individually randomized to intervention or comparator arms in a 1:1 allocation, using blocked randomization. Blocks of 4, 6 or 8 will be randomly selected to conceal allocation and render it unpredictable. The primary statistician (ACM) will generate the randomization scheme using Stata and will ensure reproducibility through setting a seed. The randomization sequence will be uploaded to the REDCap randomization module—the allocation table will not be tied at that point to individuals. Access to the allocation table will be restricted so that staff involved in recruitment, eligibility screening, enrollment, and/or outcome data collection cannot view upcoming assignments. After a participant has completed enrollment and baseline assessments, the study nurse will randomize the participant, and then receive the REDCap allocation, which will reveal only that participant’s group assignment.

#### 5.2. Blinding

This is an unblinded trial with respect to participants, educators delivering the intervention, and study supervisors, as the nature of the intervention (food supplementation and behavioral counseling) prevents blinding. However, study nurses who conduct visits for outcome data collection will be blinded. To maintain blinding, participants will be instructed not to disclose their group assignment during evaluation visits, and data collection materials will not contain information on allocation. The study statistician, after generating the sequence and uploading it to REDCap will not have further access to the sequence. The statistician will remain blinded throughout the trial until the primary analysis dataset is locked, receiving data with “Arm A” or “Arm B” assigned by the site coordinator rather than intervention or control.

### 6. Analysis Populations

Definitional note: A dyad refers to one randomized pregnant woman and her index child, both of whom contribute outcomes (woman to CP-2 and index child to CP-1). Exclusions described below may apply to one member of a dyad (e.g., the woman is excluded from CP-2, but her child is retained in CP-1) or to both dyad members (“symmetric”).

| **Population** | **Definition** | **Applies to** | **Used For** |
| --- | --- | --- | --- |
| Intent-to-Treat (ITT) | All dyads, regardless of adherence, subsequent pregnancy, or dropout. | Both women and children | Primary: CP-1; Sensitivity: CP-2 |
| Modified Intent-to-Treat (mITT) | Women who did not have a confirmed subsequent pregnancy prior to the 12-month follow-up assessment. | Women only | Primary: CP-2 |
| Symmetric mITT | Children of women who did not have a confirmed subsequent pregnancy prior to the 12-month follow-up assessment (matching the exclusion applied to women in the CP-2 primary). | Children only | Sensitivity (SA-1): CP-1 |
| Per-Protocol (CP-1) | Children in the ITT population who met all eligibility criteria, had no major protocol deviations, and whose dyad achieved the pre-specified minimum adherence thresholds (see Section 7.5). This is an as-observed subpopulation, not a principal stratum; estimates from this population are descriptive and subject to selection bias from post-randomization conditioning. | Children only | Sensitivity (SA-4): CP-1 |
| Per-Protocol (CP-2) | Women in the mITT population (i.e., women without a confirmed subsequent pregnancy prior to the 12-month follow-up assessment) who met all eligibility criteria, had no major protocol deviations, and achieved the pre-specified minimum adherence thresholds (see Section 7.5). This is an as-observed subpopulation, not a principal stratum; estimates from this population are descriptive and subject to selection bias from post-randomization conditioning. | Women only | Sensitivity (SA-4): CP-2 |
| Safety Population | All dyads who received any component of the intervention. | Both women and children | Safety analyses |

NOTE: The mITT sensitivity analysis is applied symmetrically to both co-primary outcomes (i.e., the same dyads are excluded for both CP-1 and CP-2). Asymmetric exclusion across the two co-primary endpoints is pre-specified as a primary analysis.

### 7. Statistical Analyses

#### 7.1. General Principles

- All hypothesis tests are two-sided with α = 0.05 unless otherwise specified.
- Confidence intervals are presented at the 95% level throughout.
- All analyses will be conducted using Stata version 19 or higher or R 4.5 or higher
- The biostatistician conducting primary analyses will remain blinded until the until the primary analysis dataset is locked.
- Continuous variables will be summarized as mean (SD) or median (IQR) depending on distribution. Categorical variables as n (%).
- Missing outcome data will be handled by multiple imputation as needed (see Section 8).

#### 7.2. Baseline Comparability

Baseline characteristics will be tabulated by randomization arm for all randomized dyads. No formal hypothesis testing will be conducted on baseline variables, as any observed differences are due to chance given proper randomization. At a minimum, the following will be reported:

| **Domain** | **Variables** |
| --- | --- |
| Maternal demographics | Age, parity, education, socioeconomic status, ethnicity, language preference, marital status, household wealth, gestational age at enrollment |
| Maternal anthropometrics | Height, weight gestational age at enrollment |
| Infant characteristics | Sex, gestational age at birth, birth weight |

#### 7.3. Primary Analyses of Co-Primary Endpoints

Both CP-1 and CP-2 must achieve p < 0.05 (two-sided) for the trial to be declared successful. Each is tested independently at the full α. No Bonferroni or other correction is applied between co-primary endpoints under the AND framework.

##### 7.3.1. Infant Endpoint (CP-1)

- Model**:** Linear regression of child LAZ at aged 12 months on treatment arm indicator with additional pre-specified covariates listed in Section 8.3.
- Estimand**:** Adjusted mean difference in LAZ at 12 months (intervention minus comparator) with 95% CI and two-sided p-value.
- Population**:** Full ITT — all randomized dyads including those with subsequent maternal pregnancy. See section 6 Subsequent pregnancy in the mother is not expected to directly determine the index child's LAZ at 12 months. Dyads with subsequent pregnancy are retained in the ITT analysis.

##### 7.3.2. Maternal Endpoint (CP-2)

- Model: Linear regression of maternal weight at 12 months postpartum on treatment arm indicator with additional pre-specified covariates listed in Section 8.3.
- Estimand: Adjusted mean difference in 12-month weight (kg) between arms. A negative estimate indicates lower 12-month weight in the intervention arm, consistent with reduced postpartum weight retention. 95% CI and two-sided p-value reported.95% CI and two-sided p-value reported.
- Population: modified ITT — all randomized women except those with subsequent pregnancy before 12 months. Handling of subsequent maternal pregnancy is specified in See Section 3.4 (estimand strategy) and Section 6 (analysis populations).

### 8. Secondary Analysis of Co-Primary Endpoints (Joint / Multivariate Analysis)

We will jointly model CP-1 and CP-2 simultaneously to account for within-dyad correlation and assess robustness of primary findings using MANOVA or GEE with exchangeable or unstructured working correlation matrix. The joint model population will only be performed on complete dyads; all randomized dyads except those in which the mother had a subsequent pregnancy or the child was deceased prior to 12 months of age.

#### 8.1. Adherence and Per-Protocol Analyses

Criteria for inclusion in the per-protocol population:

- Eligibility: Met all study eligibility criteria at enrollment.
- Protocol compliance: No major protocol deviations identified within protocol deviation log.
- Adherence: Receipt of ≥75% of scheduled monthly food ration deliveries (food supplementation) and completion of ≥75% of scheduled monthly educator home visits (behavioral counseling), each measured from enrollment through 12 months postpartum.

#### 8.2. Secondary Endpoint Analyses

Secondary endpoints (Section 3.2) will be analyzed using appropriate regression models consistent with the primary analysis framework. Results will be presented as point estimates, 95% CIs, and p-values.

#### 8.3. Covariate Adjustment

The following covariates will be included in all primary regression models:

| **Covariate** | **Rationale** | **Applied To** |
| --- | --- | --- |
| Randomization arm | Treatment indicator (primary predictor) | CP-1 and CP-2 |
| Maternal age at enrollment | Associated with both child growth and postpartum weight retention; adolescent pregnancies carry additional stunting and NCD risk | CP-1 and CP-2 |
| Maternal height cm | Strong predictor of child linear growth reflecting intergenerational nutritional status | CP-1 |
| Gestational age at enrollment | Associated with postpartum weight retention | CP-2 |
| Gestational age at birth | Associated with both child growth and postpartum weight retention | CP-1 |
| Parity | Higher parity is associated with lower child linear growth in low-resource settings | CP-1 |
| Baseline maternal weight | Strongest predictor of 12-month postpartum weight; also controls for regression to mean | CP-2 |
| Sex of child | Possible interaction effect as biologic sex affects growth; sex-specific differences in early linear growth are well established | CP-1 |

No covariates will be added to models post-hoc based on observed data or to achieve statistical significance. This list is final.

#### 8.4. Subgroup Analyses

All subgroup analyses are exploratory and hypothesis-generating. They are not powered to detect subgroup effects. Results should be interpreted with caution given the number of tests performed.

The following pre-specified subgroup analyses will be conducted for each co-primary endpoint by including a treatment-by-subgroup interaction term in the primary model:

| **Subgroup Variable** | **Levels** | **Scientific Rationale** |
| --- | --- | --- |
| Infant sex | Male / Female | Known sex differences in nutritional outcomes |
| Parity | 0, 1, 2 or more | Women may retain weight after each pregnancy; more experienced mothers may have different fidelity of the intervention |
| Socio-economic status | Quintiles | Both nutritional and behavioral interventions may be influenced by household wealth. Fidelity to the intervention or dropout may be related as well. |

### 9. Pre-Specified Sensitivity Analyses of co-primary endpoints

| **ID** | **Analysis** | **Endpoints** | **Purpose** |
| --- | --- | --- | --- |
| SA-1a | Exclude from the CP-1 analysis children whose mothers had a subsequent pregnancy before 12-month follow-up. This aligns the CP-1 analysis population with the CP-2 primary analysis population by excluding the same dyads from both endpoints. This corresponds to the symmetric mITT population. | CP-1 | Assess robustness of the primary CP-1 estimate when excluding these children |
| SA-1b | Include in the CP-2 analysis women with a confirmed subsequent pregnancy prior to the 12-month follow-up assessment (full ITT population). This corresponds to a treatment-policy estimand for CP-2 with respect to the subsequent-pregnancy intercurrent event. | CP-2 | Assess robustness of the principal-stratum CP-2 estimate. |
| SA-2 | Complete case analysis (no imputation). Compare to primary imputed analysis. | CP-1 and CP-2 | Assess sensitivity to missing data imputation assumptions |
| SA-3 | MNAR sensitivity: Pattern-mixture model or Tipping Point analysis assuming missing outcomes are systematically worse (or better) than observed. | CP-1 and CP-2 | Assess robustness under missing-not-at-random assumption |
| SA-4 | Per-protocol analysis restricted to dyads meeting pre-specified adherence thresholds for the relevant component. | CP-1 and CP-2 (separately, using component-specific PP population) | Describe the treatment-effect estimate among adherers; compare to the primary ITT/mITT estimates to assess sensitivity to non-adherence. |
| SA-5 | Analyses for all of the above with augmented product terms for sex and main analysis factors | CP-1 | Assess influence of biologic sex of child on the co-primary outcome of LAZ score at 12 months. |
| SA-6 | Fully adjusted models for all of the above including maternal education and maternal continuous SES score | CP-1 and CP-2 | Assess robustness of primary estimates to further adjustment for socioeconomic covariates. |
| SA-7 | Unadjusted analysis | CP-1 and CP-2 | Assess influence of covariate adjustment on point estimates |

### 10. Missing Data

#### 10.1. Expected Missing Data Sources

- Withdrawal
- Loss to follow-up (relocation, inability to contact)
- Missed study visits / partial data collection at a visit
- Subsequent pregnancy resulting in modified measurement context (note: child remains in ITT)
- Death (outcome data will not be imputed, but baseline data will inform imputation models)

#### 10.2. Missing Data Assumptions and Primary Approach

**Primary assumption:** Missing at Random (MAR) — missingness depends on observed data but not on the unobserved outcome value after conditioning on observed covariates.

**Primary method:** Multiple Imputation with a minimum of [M = 10] imputed datasets. Variables to be imputed will depend on missingness patterns. The imputation model will include (if necessary): treatment arm, both co-primary outcomes at baseline and 12 months, all covariates in the primary analysis model, auxiliary variables associated with missingness (e.g., number of visits attended, baseline health status). Rubin's rules will be applied to pool estimates across imputed datasets.

Imputation will be conducted separately for each co-primary endpoint

**Alternative method**: Full Information Maximum Likelihood using Stata’s SEM commands or R. We will use Maximum Likelihood if only the outcome data are missing. In the event that primary predictor data are missing, we will assess the amount of missing data. If 10% or less, we will handle missing data through Full Information Maximum Likelihood using Stata’s SEM commands. Otherwise, we will use multiple imputation as described above.

Other statistical packages may be used for the analyses but if the above prespecified tools are not used, this will be specified in the final report.

#### 10.3. Sensitivity for Missing Data

SA-2 (complete case) and SA-3 (MNAR via pattern-mixture or tipping point) are pre-specified in Section 8. Results will be compared to primary imputed analyses and discrepancies discussed in the study report.

### 11. Interim Analyses and Data Monitoring

No formal interim analyses are planned.

### 12. Safety Analyses

While the trial is of not more than minimum risk and no adverse events related to trial participation are expected, safety analyses will be conducted in the Safety Population (all dyads receiving any intervention). The following will be summarized descriptively by arm:

| **Safety Domain** | **Summary Approach** |
| --- | --- |
| Adverse events (AEs) | Number and % of dyads with ≥1 AE; events by system organ class and preferred term |
| Serious adverse events (SAEs) | Listed individually; n (%) by arm |
| AEs leading to discontinuation | n (%) by arm |

No formal hypothesis testing is planned for safety endpoints. Formal causal inference on safety is beyond the scope of this trial. All safety data will be presented to the DSMB per the DSMB Charter.

25.	World Health Organization. Guideline on haemoglobin cutoffs to define anaemia in individuals and populations. Geneva: World Health Organization, 2024.

26.	Ministerio de Salud Pública y Asistencia Social. Normas de Atención Integral para la Red Integrada de Servicios de Salud. Guatemala: Viceministerio de Regulación, Vigilancia y Control de la Salud, Dirección de Normatividad de Programas de Atención a las Personas, República de Guatemala, 2025.

27.	World Health Organization (WHO). Stillbirths. 2026. <https://www.who.int/maternal_child_adolescent/epidemiology/stillbirth/en/> (accessed February 11, 2026).

28.	Simondon KB, Simondon F, Delpeuch F, Cornu A. Comparative study of five growth models applied to weight data from congolese infants between birth and 13 months of age. *Am J Hum Biol* 1992; 4(3): 327-35.

29.	Valderrama CE, Marzbanrad F, Juarez M, Hall-Clifford R, Rohloff P, Clifford GD. Estimating birth weight from observed postnatal weights in a Guatemalan highland community. *Physiol Meas* 2020; 41(2): 025008.

30.	WHO Multicentre Growth Reference Study Group. WHO Child Growth Standards based on length/height, weight and age. *Acta Paediatr Suppl* 2006; 450: 76-85.

31.	World Health Organization. Indicators for assessing infant and young child feeding practices: definitions and measurement methods. 2021.
